# Reporting Rates for VAERS Death Reports Following COVID-19 Vaccination, December 14, 2020-November 17, 2021

**DOI:** 10.1101/2022.05.05.22274695

**Authors:** Brendan Day, David Menschik, Deborah Thompson, Christopher Jankosky, John Su, Pedro Moro, Craig Zinderman, Kerry Welsh, Narayan Nair

## Abstract

**Background:** Despite widely available safety information for the COVID-19 vaccines, vaccine hesitancy remains a challenge. In some cases, vaccine hesitancy may be related to concerns about the number of reports of death to the Vaccine Adverse Event Reporting System (VAERS).

**Objective:** To provide information and context about reports of death to VAERS following COVID-19 vaccination.

**Design:** Descriptive study; reporting rates for VAERS death reports.

**Setting:** United States; December 14, 2020, to November 17, 2021.

**Participants:** COVID-19 vaccine recipients.

**Measurements:** Reporting rates for death events per million persons vaccinated; adverse event counts; data mining signals of disproportionate reporting.

**Results:** 9,201 death events were reported for COVID-19 vaccine recipients aged five years and older (or age unknown). Reporting rates for death events increased with increasing age, and males generally had higher reporting rates than females. For death events within seven days and 42 days of vaccination, respectively, observed reporting rates were lower than the expected all-cause death rates. Reporting rates for Ad26.COV2.S vaccine were generally higher than for mRNA COVID-19 vaccines, but still lower than the expected all-cause death rates. Reported adverse events were non-specific or reflected the known leading causes of death.

**Limitations:** VAERS data are subject to several limitations such as reporting bias (underreporting and stimulated reporting), missing or inaccurate information, and lack of a control group. Reported diagnoses, including deaths, are not causally verified diagnoses.

**Conclusion:** Reporting rates for death events were lower than the expected all-cause mortality rates. Trends in reporting rates reflected known trends in background mortality rates. These findings do not suggest an association between vaccination and overall increased mortality.

**Funding Source:** No external sources of funding were used.

## Introduction

As of November 17, 2021, the Coronavirus Disease 2019 (COVID-19) pandemic had led to the deaths of 761,426 Americans (1). In December 2020, the Food and Drug Administration (FDA) issued Emergency Use Authorizations (EUAs) for the Pfizer-BioNTech (BNT162b2) and Moderna (mRNA-1273) vaccines for the prevention of COVID-19. In February 2021, Johnson & Johnson’s Janssen (Ad26.COV2.S) COVID-19 vaccine became the third vaccine available under EUA. Within a year of the first vaccines being authorized, over 239 million individuals in the United States (US) had received at least one dose of a COVID-19 vaccine (2). According to a December 2021 report by the Office of the Assistant Secretary for Planning and Evaluation (ASPE), as of July 2021, COVID-19 vaccinations in the US were associated with estimated reductions of approximately 213,000 deaths and 1.38 million hospitalizations (3). More recently, a group of university-based researchers arrived at similar conclusions; in a research letter published in January 2022, Vilches et al. estimated that the US COVID-19 vaccination campaign through June 2021 had saved over 240,000 lives and prevented over 1.1 million hospitalizations (4).

Despite the large number of people who have received these vaccines safely, and the widespread availability of information regarding the safety and efficacy of the COVID-19 vaccines, vaccine hesitancy remains a challenge (5). In some cases, vaccine hesitancy may be related to misleading claims about the number of reports of death following vaccination to the Vaccine Adverse Event Reporting System (VAERS). Specifically, some media reports have incorrectly described the number of death reports in VAERS as a “death toll,” which falsely implies that these deaths were necessarily caused by vaccination (6-9).

VAERS is a national vaccine safety surveillance program that was established in 1990 and is co-managed by the Centers for Disease Control and Prevention (CDC) and FDA (10). VAERS serves as a hypothesis-generating, early warning system that scientists at FDA and CDC use to detect unusual or unexpected patterns of adverse events (also known as “safety signals”) that warrant further investigation (11, 12). As a passive reporting system, VAERS relies on individuals reporting adverse events after vaccination. Anyone can submit reports to VAERS, including patients, family members, healthcare providers, and vaccine manufacturers (13). No proof that the event was caused by the vaccine is required for VAERS to accept the report (14). Serious and unexpected medical events (such as death) are probably more likely to be reported than minor events, even if they may be coincidental and related to other causes (14). Reports of adverse events, including death, do not necessarily mean that the reported problem was caused by a vaccine. This is especially true for VAERS reports for COVID-19 vaccines, since healthcare providers who administer COVID-19 vaccines are legally required to report to VAERS any deaths following vaccination, regardless of causality (15). (For mandatory reporting requirements, see each COVID-19 vaccine “Fact Sheet for Healthcare Providers Administering Vaccine”) (16-18). For these reasons, interpreting VAERS data without an understanding of its purpose, strengths, and limitations can lead to erroneous conclusions about cause and effect (19).

The objective of this paper is to provide information and context about reports of death to VAERS following COVID-19 vaccination by 1) assessing reporting rates for deaths reported after COVID-19 vaccination and comparing these reporting rates to expected background rates of deaths from all causes in the general population, and 2) evaluating the most common adverse events in VAERS death reports for COVID-19 vaccines and comparing these reported adverse events to the known leading causes of death in the US.

## Methods

### Content and Processing of VAERS Reports

The VAERS reporting form includes a description of the adverse event(s), the name(s) of the vaccine(s) received, the dates of vaccination and adverse event onset, the result or outcome of the adverse event(s) (including whether the patient died), as well as medical and demographic information for the patient (20). In addition, reporters can submit supporting documents such as vaccination cards, medical records, death certificates, or autopsy reports. As part of routine processing following submission, VAERS staff with expertise in coding case report information review reports and assign medical terms for adverse events using an international standardized coding system called MedDRA (Medical Dictionary for Regulatory Activities) (21). Each reported adverse event is linked to a MedDRA term called a Preferred Term. MedDRA terms are not confirmed medical diagnoses, but rather serve as a coding scheme to classify and encode adverse event information in VAERS reports (19). For US reports of death following COVID-19 vaccination, VAERS staff request and review associated medical records, including hospital discharge summaries, test results, autopsy reports, and death certificates (22).

### VAERS Strengths and Limitations

Strengths of VAERS include the ability to rapidly detect potential safety signals, inclusion of reports from the entire US, and the potential to detect rare adverse events. VAERS data are also made available online to the public (https://vaers.hhs.gov/data.html), which affords an important level of transparency. The limitations of VAERS include reporting bias (including underreporting of adverse events, especially mild events) and stimulated reporting, which is increased reporting in response to media attention or heightened public awareness. There may be missing or inaccurate information in reports, and reported diagnoses are not verified. The lack of an unvaccinated control group prevents comparison of rates between exposed and unexposed individuals. Different limitations, such as underreporting and the lack of a denominator, can prevent calculation of how often an adverse event occurs in a population (11, 19). Because of these limitations, VAERS data alone are generally insufficient for determining causality for a given adverse event after vaccination (19, 23).

### VAERS Reporting Requirements and Death Reports

As with licensed non-COVID-19 vaccines, anyone can submit VAERS reports for COVID-19 vaccines available under EUA. However, unlike non-COVID-19 vaccines, healthcare providers administering COVID-19 vaccines under EUA are legally required to report to VAERS *all* deaths after vaccination, irrespective of attribution to vaccination. They are also required to report to VAERS any cases of COVID-19 that occur after vaccination and result in death (13, 16-18). This differs from non-COVID-19 vaccines, for which healthcare providers are only required to report deaths if they meet specific criteria on the “VAERS Table of Reportable Events Following Vaccination” (24).

Deaths are identified in VAERS reports when “Patient died” is selected as a result or outcome of the adverse event(s) in Box 21 of the VAERS reporting form (20). Each VAERS report has a “primary” or initial report and may also have several subsequent or follow-up reports (12). For example, a patient’s caregiver may submit a primary report and a healthcare professional may subsequently submit a separate report for the same deceased individual. Due to privacy protections, publicly available VAERS data contain only the primary report, with sensitive patient information removed (19). Conversely, the federal VAERS database contains both primary and subsequent reports. Therefore, the federal VAERS database can contain multiple reports for the same deceased person. To address this redundancy, we use the term “death event” to describe an individual death, which may be reported in more than one VAERS death report. The term “death event” does not imply a verified or confirmed diagnosis, only that that an individual was reported to have died after being vaccinated.

### Analysis of US VAERS Death Reports

For our main analysis, we calculated reporting rates for death events reported any time after COVID-19 vaccination in US VAERS reports received between December 14, 2020, and November 17, 2021, and we compared these reporting rates to expected background rates of death from all causes in the general population. COVID-19 vaccines included in our analysis were the three COVID-19 vaccines available under EUAs during the study period: BNT162b2 vaccine (initially authorized on December 11, 2020), mRNA-1273 vaccine (initially authorized on December 18, 2020), and Ad26.COV2.S vaccine (initially authorized on February 27, 2021). Reporting rates are calculated by dividing the number of reported events (i.e., deaths) by the number of people exposed to the vaccine (i.e., people vaccinated with at least one dose of a COVID-19 vaccine). Reporting rates using VAERS data are subject to the same limitations mentioned above and must be interpreted with caution. Because of these limitations, reporting rates should not be considered equivalent to incidence rates. Nonetheless, they serve as a useful pharmacovigilance tool in detecting safety signals that may warrant further investigation.

For example, a reporting rate that is higher than the background rate may indicate that the true incidence rate is sufficiently high to be a concern, and further evaluation to assess for a causal relationship to the vaccine may be necessary. Conversely, a reporting rate that is below the background rate would be expected if there were no relationship between the adverse event and the vaccine. However, due to the limitations of reporting rates, the vaccine could still be associated with an increased risk of the adverse event (25).

To determine the number of death events reported in VAERS from US sources (i.e., US death events) for the COVID-19 vaccines, on November 17, 2021, we queried the VAERS database using a standard set of consolidation rules (Table S1). According to these rules, we included any US death events where at least one of the associated VAERS reports had an outcome of “Patient died.” All foreign source VAERS reports were excluded. We included all reported US death events and did not perform causality assessments for individual cases. We obtained vaccine administration data from the CDC regarding the number of individuals in the US (by age and by sex) vaccinated with at least one dose of a COVID-19 vaccine. We then determined characteristics (age and sex of vaccine recipient) and reporting rates (per million persons vaccinated with at least one dose of a COVID-19 vaccine) for death events after COVID-19 vaccination. Because the EUA for the BNT162b2 vaccine allows for vaccination of individuals as young as five years old, we excluded individuals with a reported age of under 5 years from all of our reporting rate calculations.

For death events with a non-negative onset interval (i.e., a known death date that occurred on or after a known vaccination date), we compared reporting rates for death events within seven days (days 0-7, inclusive) and within 42 days (days 0-42, inclusive) after COVID-19 vaccination with expected all-cause death rates as determined by Abara et al. using data from the National Vital Statistics System (26). These expected all-cause death rates show how many deaths in the US population would be expected to occur coincidentally (from causes unrelated to vaccination) within seven and 42 days of vaccination, respectively.

To evaluate adverse events reported in US VAERS death reports, we queried the VAERS database for the 25 most common MedDRA Preferred Terms (PTs) in US VAERS death reports and compared these PTs to the leading causes of death in the US from 2015-2020 as reported by the National Center for Health Statistics (27).

Lastly, we performed Empirical Bayesian (EB) data mining to evaluate for signals of disproportionate reporting among US VAERS death reports for COVID-19 vaccines (28). Data mining covers the entire post-authorization period for each vaccine from the initial authorization through the data lock date listed. The background database contains VAERS reports since 1990. Adjusting for age, sex, and year, EB data mining generates ratios of observed-to-expected counts [Empirical Bayes Geometric Mean (EBGM)] for vaccine-adverse event combinations. Results greater than two for the lower bound of the 90% confidence interval for the EBGM (i.e., EB05 > 2) are considered signals of disproportionate reporting; this indicates, with a high degree of confidence, that a given vaccine-adverse event is being reported at least twice as frequently as expected if there were no association (28, 29). Data mining findings are subject to a number of limitations and do not imply causality; rather, they should be regarded as “hypothesis-generating” (28).

### Ethics

Because VAERS is a vaccine safety surveillance program, this study was exempt from Institutional Review Board review. Activities were reviewed by the FDA and CDC and conducted in accordance with applicable federal law and FDA and CDC policy.

## Results

### Characteristics and Reporting Rates for Death Events

As of November 17, 2021, there were 9,790 US death reports representing 9,205 death events for all COVID-19 vaccines. After restricting to individuals aged 5 years and older or with unknown age, there were 9,201 death events reported for all COVID-19 vaccines (Table 1). For each vaccine, the proportion of death events was higher for males than females. With regard to age, the proportion of death events for the mRNA COVID-19 vaccines increased with increasing age and peaked in the age group 75-84 years. For the Ad26.COV2.S vaccine, the proportion of death events peaked in the age groups 55-64 years and 65-74 years. The Ad26.COV2.S vaccine also had the highest proportion of unknown age for death events (24.9%).

**Table 1:** Characteristics and reporting rates (death events per million vaccinated persons) for US death events reported to Vaccine Adverse Event Reporting System (VAERS) among COVID-19 vaccine recipients*—Reports received between December 14, 2020-November 17, 2021

|  | All COVID-19 vaccines |  | BNT162b2 vaccine |  | mRNA-1273 vaccine |  | Ad26.COV2.S vaccine |  |
| --- | --- | --- | --- | --- | --- | --- | --- | --- |
|  | (n=9,201)*†‡ |  | (n=4,207) |  | (n=3,877) |  | (n=1,056) |  |
|  | n (%) | Reporting rate§ | n (%) | Reporting rate§ | n (%) | Reporting rate§ | n (%) | Reporting rate§ |
| Sex |  |  |  |  |  |  |  |  |
| Female | 3,854 (41.9) | 35.2 | 1,802 (42.8) | 28.6 | 1,608 (41.5) | 40.1 | 428 (40.5) | 68.1 |
| Male | 4,949 (53.8) | 49.7 | 2,183 (51.9) | 39.2 | 2,197 (56.7) | 61.2 | 529 (50.1) | 67.0 |
| Unknown | 398 (4.3) | 189.7 | 222 (5.3) | 221.6 | 72 (1.9) | 79.5 | 99 (9.4) | 525.3 |
| Age (years) |  |  |  |  |  |  |  |  |
| 5-14 | 13 (0.1) | 1.4 | 12 (0.3) | 1.3 | 1 (0.03) | 91.4¶ | 0 (0) | 0.0 |
| 15-24 | 88 (1.0) | 3.4 | 54 (1.3) | 3.0 | 20 (0.5) | 3.2 | 13 (1.2) | 8.2 |
| 25-34 | 146 (1.6) | 5.0 | 69 (1.6) | 4.1 | 60 (1.5) | 6.0 | 17 (1.6) | 7.0 |
| 35-44 | 284 (3.1) | 9.5 | 117 (2.8) | 7.0 | 114 (2.9) | 10.7 | 52 (4.9) | 20.1 |
| 45-54 | 485 (5.3) | 15.8 | 189 (4.5) | 11.5 | 206 (5.3) | 18.0 | 85 (8.0) | 31.3 |
| 55-64 | 1,146 (12.5) | 32.8 | 486 (11.6) | 27.4 | 462 (11.9) | 32.6 | 190 (18.0) | 64.0 |
| 65-74 | 2,001 (21.7) | 65.8 | 891 (21.2) | 61.1 | 905 (23.3) | 62.8 | 190 (18.0) | 137.8 |
| 75-84 | 2,199 (23.9) | 137.6 | 1,013 (24.1) | 131.9 | 1,032 (26.6) | 133.3 | 140 (13.3) | 267.1 |
| ≥85 | 2,034 (22.1) | 426.0 | 996 (23.7) | 420.0 | 921 (23.8) | 413.5 | 106 (10.0) | 642.3 |
| Unknown | 805 (8.7) | 29,149.8 | 380 (9.0) | 33,173.3 | 156 (4.0) | 11,359.5 | 263 (24.9) | 114,447.3 |
| Total | 9,201 (100) | 43.6 | 4,207 (100) | 35.2 | 3,877 (100) | 50.4 | 1,056 (100) | 73.5 |

\* Excludes individuals with known age less than 5 years old. Includes individuals aged 5 years and older or with unknown age. Includes 85 death events with unknown COVID-19 vaccine name.
† All COVID-19 vaccines includes 24 deaths with more than one COVID-19 vaccine reported. BNT162b2 vaccine includes 14 deaths with both the BNT162b2 vaccine and another COVID-19 vaccine reported. mRNA-1273 vaccine includes 16 deaths with the both mRNA-1273 vaccine and another COVID-19 vaccine reported. Ad26.COV2.S vaccine include 8 deaths with both the Ad26.COV2.S vaccine and another COVID-19 vaccine reported.
‡ At the time of data cutoff (November 17, 2021), the BNT162b2 vaccine was authorized for use in individuals aged 5 years and older, the mRNA-1273 vaccine was authorized for use in individuals aged 18 years and older, and the Ad26.COV2.S vaccine was authorized for use in adults aged 18 years and older.
§ Reporting rate for death events per million persons vaccinated with at least one dose of a COVID-19 vaccine.
II Vaccine administration data: Includes vaccines administered between 12/14/2020 and 11/17/21 as reported to the CDC as of 6 AM 11/29/2021; Excludes vaccine refusals and vaccines administered by DoS (Department of State); Includes U.S. territories; Does not include vaccine administrations reported by Texas. Texas provides aggregate dose count data to CDC. Therefore, specific information at the individual dose level is not available. Numbers provided may still include vaccine administrations for Texas residents due to vaccines administered out-of-state or by federal entities; Does not include vaccine administrations reported by Idaho for recipients under 18 years. Idaho provides vaccine data only for vaccine recipients who are 18 years and over, in line with state laws. Numbers provided may still include vaccine administrations for Idaho residents under 18 years due to vaccines administered out-of-state or by federal entities; All COVID-19 vaccines includes vaccines of Unknown type.
¶¶ Because the mRNA-1273 vaccine was not authorized for use in this age group, the denominator for this reporting rate was relatively small compared to other age groups, thus allowing the single reported death event in this age group to lead to an elevated reporting rate.

Reporting rates for death events generally followed similar trends across all three COVID-19 vaccines (Table 1). Males had higher reporting rates than females for each vaccine except for the Ad26.COV2.S vaccine, which had comparable reporting rates for males and females. However, the Ad26.COV2.S vaccine also had a higher reporting rate for death events with unknown sex than the mRNA COVID-19 vaccines. Reporting rates for each vaccine increased with increasing age, peaking in the oldest age group (≥85 years). With the exception of the youngest age group, the Ad26.COV2.S vaccine had the highest reporting rates for each age group as well as overall.

### Expected and Observed Reporting Rates for Death Events

For death events with a non-negative onset interval, the median time to death was 11 days post-vaccination (interquartile range: 2-58 days). Table 2 displays the number of expected all-cause deaths compared to the observed reporting rate for death events for COVID-19 vaccines within seven days and within 42 days of vaccination, respectively. For each vaccine, across all sex and age groups, the observed reporting rate for death events was much lower than the number of expected all-cause deaths. For all age groups combined, reporting rates for each vaccine and for all COVID-19 vaccines combined were lower than expected all-cause deaths. For all COVID-19 vaccines combined, the observed reporting rates for US death events was approximately 10 times lower than the expected all-cause death rate within seven days of vaccination and approximately 36 times lower than the expected all-cause death rate within 42 days of vaccination.

**Table 2:**
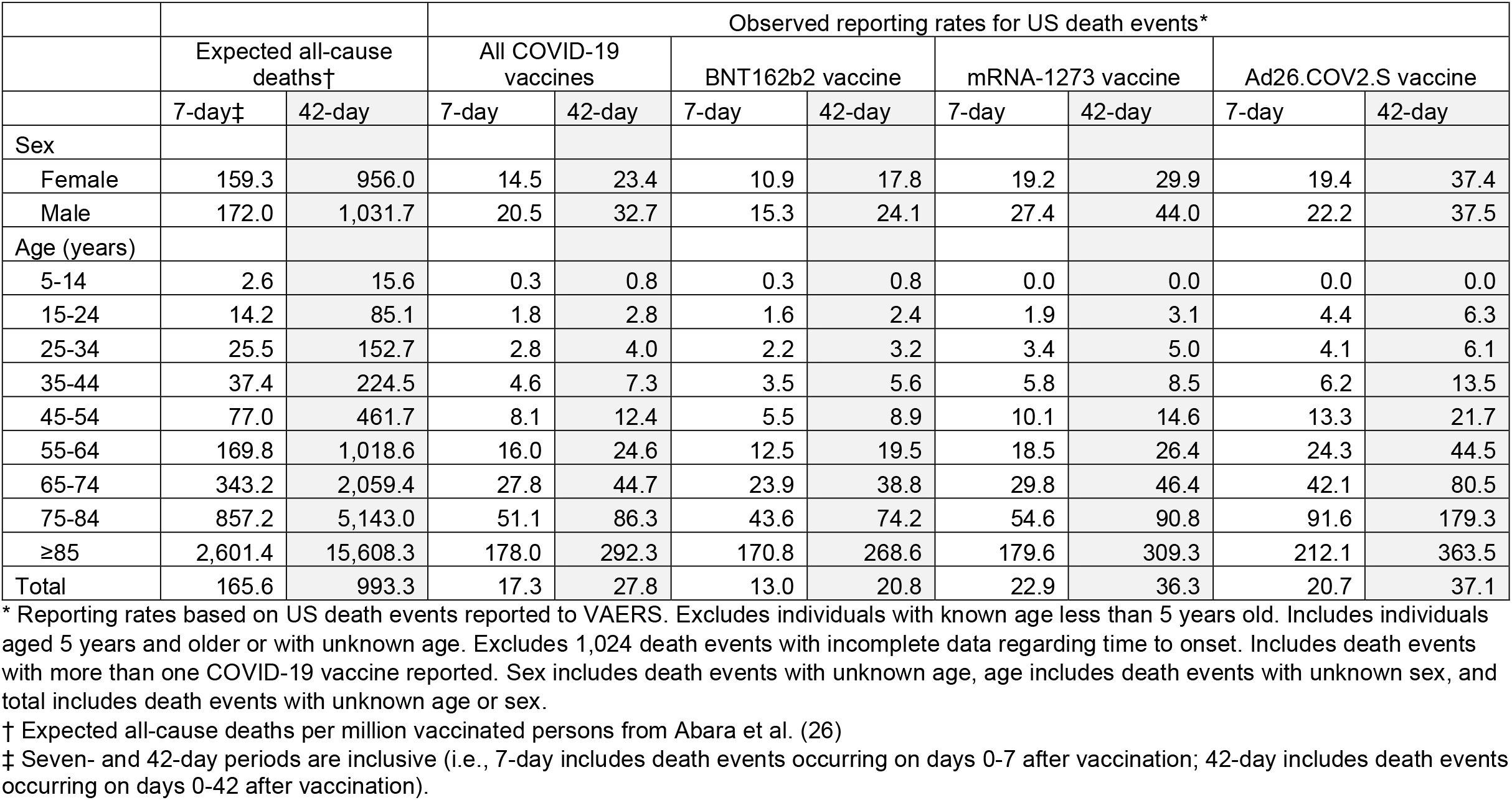
Expected rates of death per million vaccinated persons and US death events reported to VAERS per million vaccinated persons within 7 days and within 42 days of vaccination among COVID-19 vaccine recipients, by sex and age—Reports received between December 14, 2020-November 17, 2021

|  | Expected all-cause deaths† |  | Observed reporting rates for US death events* |  |  |  |  |  |  |  |
| --- | --- | --- | --- | --- | --- | --- | --- | --- | --- | --- |
|  |  |  | All COVID-19 vaccines |  | BNT162b2 vaccine |  | mRNA-1273 vaccine |  | Ad26.COV2.S vaccine |  |
|  | 7-day‡ | 42-day | 7-day | 42-day | 7-day | 42-day | 7-day | 42-day | 7-day | 42-day |
| Sex |  |  |  |  |  |  |  |  |  |  |
| Female | 159.3 | 956.0 | 14.5 | 23.4 | 10.9 | 17.8 | 19.2 | 29.9 | 19.4 | 37.4 |
| Male | 172.0 | 1,031.7 | 20.5 | 32.7 | 15.3 | 24.1 | 27.4 | 44.0 | 22.2 | 37.5 |
| Age (years) |  |  |  |  |  |  |  |  |  |  |
| 5-14 | 2.6 | 15.6 | 0.3 | 0.8 | 0.3 | 0.8 | 0.0 | 0.0 | 0.0 | 0.0 |
| 15-24 | 14.2 | 85.1 | 1.8 | 2.8 | 1.6 | 2.4 | 1.9 | 3.1 | 4.4 | 6.3 |
| 25-34 | 25.5 | 152.7 | 2.8 | 4.0 | 2.2 | 3.2 | 3.4 | 5.0 | 4.1 | 6.1 |
| 35-44 | 37.4 | 224.5 | 4.6 | 7.3 | 3.5 | 5.6 | 5.8 | 8.5 | 6.2 | 13.5 |
| 45-54 | 77.0 | 461.7 | 8.1 | 12.4 | 5.5 | 8.9 | 10.1 | 14.6 | 13.3 | 21.7 |
| 55-64 | 169.8 | 1,018.6 | 16.0 | 24.6 | 12.5 | 19.5 | 18.5 | 26.4 | 24.3 | 44.5 |
| 65-74 | 343.2 | 2,059.4 | 27.8 | 44.7 | 23.9 | 38.8 | 29.8 | 46.4 | 42.1 | 80.5 |
| 75-84 | 857.2 | 5,143.0 | 51.1 | 86.3 | 43.6 | 74.2 | 54.6 | 90.8 | 91.6 | 179.3 |
| ≥85 | 2,601.4 | 15,608.3 | 178.0 | 292.3 | 170.8 | 268.6 | 179.6 | 309.3 | 212.1 | 363.5 |
| Total | 165.6 | 993.3 | 17.3 | 27.8 | 13.0 | 20.8 | 22.9 | 36.3 | 20.7 | 37.1 |

\* Reporting rates based on US death events reported to VAERS. Excludes individuals with known age less than 5 years old. Includes individuals aged 5 years and older or with unknown age. Excludes 1,024 death events with incomplete data regarding time to onset. Includes death events with more than one COVID-19 vaccine reported. Sex includes death events with unknown age, age includes death events with unknown sex, and total includes death events with unknown age or sex.
† Expected all-cause deaths per million vaccinated persons from Abara et al. (26)
‡ Seven- and 42-day periods are inclusive (i.e., 7-day includes death events occurring on days 0-7 after vaccination; 42-day includes death events occurring on days 0-42 after vaccination).

For each COVID-19 vaccine, within seven and 42 days of vaccination, males had higher reporting rates for death events than females. Reporting rates also increased with increasing age for each vaccine within both seven and 42 days of vaccination, peaking in age group ≥85 years. With the exception of age group 5-14 years, the Ad26.COV2.S vaccine had higher reporting rates for death events than the mRNA COVID-19 vaccines for every age group within both seven and 42 days of vaccination. However, these reporting rates were still well below the expected all-cause death rates within seven and 42 days of vaccination. Furthermore, the overall reporting rates for the Ad26.COV2.S vaccine within seven and 42 days of vaccination were comparable to those of the mRNA COVID-19 vaccines.

### Most Common Preferred Terms in Death Reports

The most common Preferred Terms (PTs) among US death reports for all COVID-19 vaccines combined are displayed in Table 3. The majority of the 25 most common PTs were non-specific signs, symptoms, or investigations. PTs reflecting identifiable diseases or conditions (e.g., COVID-19 or pneumonia) were similar to the leading causes of death in 2020 as provisionally reported by the National Center for Health Statistics (27).

**Table 3:**
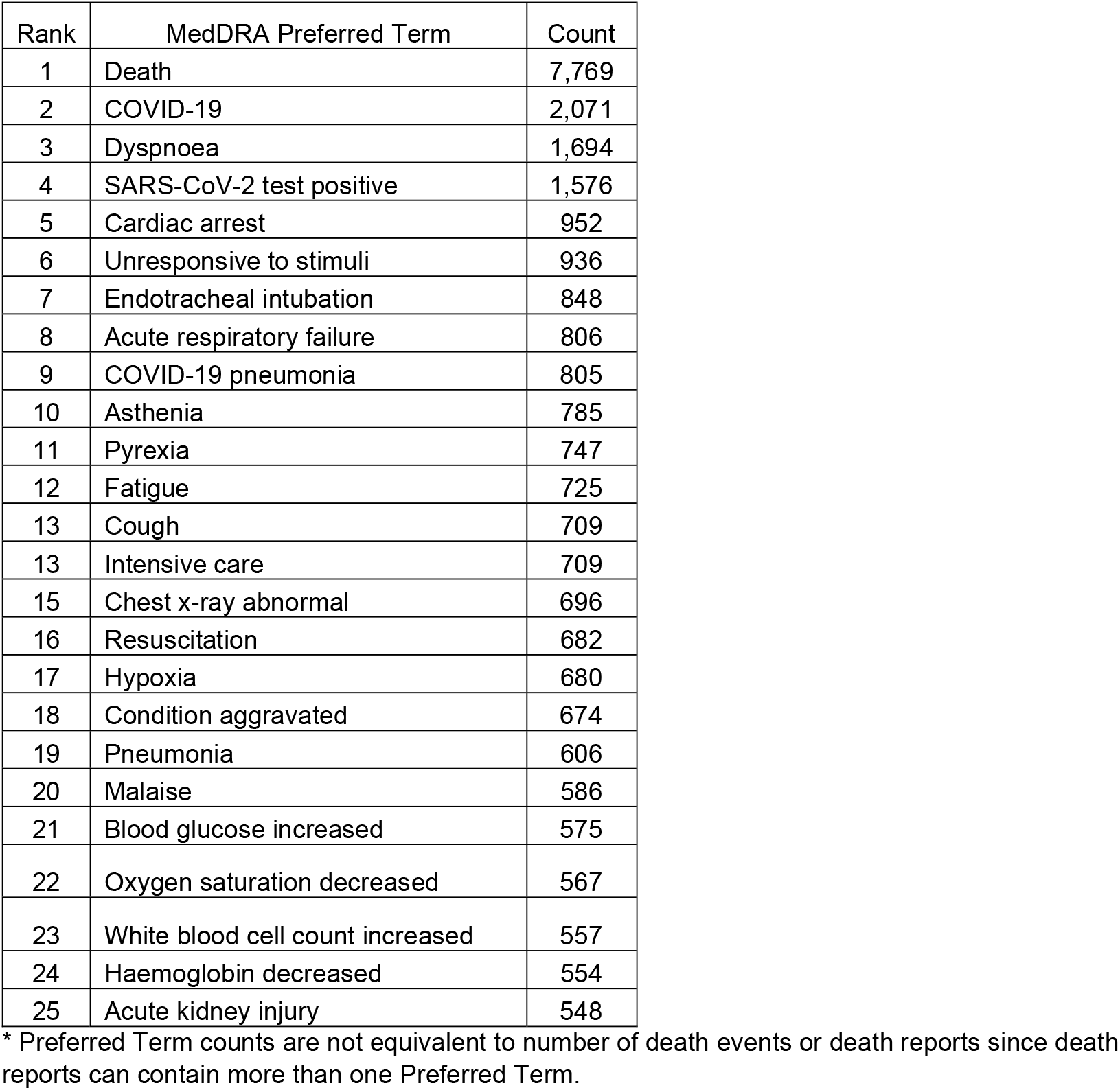
Most common Preferred Terms in US VAERS death reports for COVID-19 vaccines, all ages—Reports completed between December 14, 2020-November 17, 2021.

### Data Mining

Data mining performed on November 17, 2021 (with a data lock point of November 12, 2021), revealed one signal of disproportionate reporting (EB05>2) for US VAERS death reports. The Ad26.COV2.S vaccine had an EB05 of 2.52 for the Preferred Term “Vaccination Failure.”

## Discussion

Our analysis revealed that reporting rates for death events among COVID-19 vaccine recipients were substantially lower than expected all-cause mortality rates for each COVID-19 vaccine, both overall and across all age and sex groups (Table 2). Trends in reporting rates for death events generally reflected known trends in background death rates (i.e., increasing death rates with increasing age, males with higher death rates than females). Likewise, MedDRA PTs in death reports (Table 3) were either non-specific or reflective of the known leading causes of death in 2020 and the five years leading up to the pandemic (27). These findings do not suggest an association between vaccination and overall increased mortality.

We observed some differences in findings between the mRNA COVID-19 vaccines and the Ad26.COV2.S vaccine. Specifically, with the exception of the youngest age group (5-14 years), the Ad26.COV2.S vaccine overall had higher reporting rates for death events for each age group (Table 1) as well as higher reporting rates for death events within seven days and within 42 days postvaccination for each age group (Table 2). Lastly, the Ad26.COV2.S vaccine was the only COVID-19 vaccine with a positive data mining signal for US VAERS death reports (“Vaccination failure”). Possible explanations for these findings include the following: 1) confounding by differences in patient-specific characteristics (e.g., underlying comorbidities or health behaviors that differ between vaccinated populations), 2) differences in reporting behavior (i.e., stimulated reporting) for recipients of the Ad26.COV2.S vaccine compared to the mRNA COVID-19 vaccines, or 3) differences in real-world effectiveness of the Ad26.COV2.S vaccine compared to the mRNA COVID-19 vaccines (30, 31). Despite these observed differences, the reporting rates for death events after the Ad26.COV2.S vaccine were still considerably less than the expected all-cause death rates.

Our study has several strengths. First, because anyone can submit reports to VAERS, this data set captures a broad diversity of demographics. Second, because VAERS triaging prioritizes US VAERS death reports (typically processed within one business day), this enables timely review of such events (22). Third, by using consolidated VAERS data, we were able to identify individual deaths and avoid overcounting deaths in situations where two or more reporters submitted separate death reports for the same deceased individual. Finally, in calculating reporting rates for death events, we used the number of individuals vaccinated with at least one dose of a COVID-19 vaccine as the denominator, rather than the number of doses administered. This resulted in more conservative estimates (i.e., higher reporting rates for death events) and allowed for comparison between vaccines, regardless of differences in dose series (i.e., two-dose versus one-dose).

Our study also has limitations. First, VAERS reports can have conflicting or missing information, which could impact the results. For example, for one of our analyses (Table 2), 1,024 death events were excluded due to invalid or missing information regarding time to onset. However, we aimed to reduce the effect of missing or conflicting information in VAERS reports by using a standard set of consolidation rules (Table S1) to capture death events and their characteristics (age, sex, and time to onset); even so, these rules could yield inaccurate values. For example, age may be captured inaccurately if a healthcare provider reports the age inaccurately, despite a family member separately reporting the correct age. Second, although this analysis demonstrates low reporting rates of death after vaccination overall, these data do not preclude causal attribution of the vaccine for verified safety signals of specific adverse events. For example, an earlier review of VAERS reports for Ad26.COV2.S vaccine identified a safety signal for thrombosis with thrombocytopenia syndrome (TTS), a rare and serious adverse event that causes blood clots with low platelets. Further evaluation of this safety signal confirmed a causal relationship, and as of December 16, 2021, nine deaths reported in VAERS had been directly attributed to TTS following Ad26.COV2.S vaccine (32).

Another potential limitation includes underreporting in VAERS. Underreporting is a well-known limitation of all passive surveillance systems and reporting sensitivities for various adverse events following immunization can vary widely (33-35). We are not aware of peer-reviewed published studies that address spontaneous adverse event reporting in the context of mandatory EUA and CDC reporting requirements. However, a previous publication using data from before the COVID-19 pandemic estimated the reporting sensitivity of VAERS for anaphylaxis and Guillain-Barré syndrome (GBS) for several non-COVID-19 vaccines, including both the seasonal influenza vaccine and the 2009 H1N1 inactivated pandemic influenza vaccine. The reporting sensitivity for the seasonal influenza vaccine was 13% for anaphylaxis and 12% for GBS. Conversely, the reporting sensitivity for the 2009 H1N1 inactivated pandemic influenza vaccine was 76% for anaphylaxis and 55% for GBS. These findings suggest that underreporting in VAERS may be mitigated in the setting of a pandemic where there is a heavy emphasis on vaccine safety. Furthermore, the COVID-19 vaccine program has the unique EUA and CDC requirement of mandatory reporting of deaths (15-18), which has likely increased capture of death reports. Lastly, data mining has several limitations, including: 1) disproportionality scores for COVID-19 vaccines can be attenuated due to a relatively high number of COVID-19 vaccine reports in the background, and 2) the absence of a statistical signal does not rule out a possible corresponding adverse event (29).

Our results are consistent with other studies, which do not show an association between COVID-19 vaccination and increased mortality. In a published article using data from approximately 11 million individuals enrolled in seven Vaccine Safety Datalink (VSD) sites, a matched cohort analysis found that COVID-19 vaccine recipients had consistently lower mortality rates than unvaccinated individuals across each of the three available COVID-19 vaccines, including after both first and second doses of the mRNA COVID-19 vaccines (36). Another cohort study, looking at nursing home residents, similarly found that individuals who received the BNT162b2 vaccine or the mRNA-1273 vaccine had lower all-cause mortality than unvaccinated residents (37). Lastly, a recently published analysis of VAERS reports through June 14, 2021, for the mRNA COVID-19 vaccines found no unusual patterns in cause of death among death reports received. Furthermore, reporting rates for death were higher in older age groups, consistent with age-specific mortality in the general adult population (38).

In conclusion, our analysis of VAERS data for COVID-19 vaccines found that reporting rates for death events were much lower than expected all-cause mortality rates, and trends in reporting rates reflected known trends in background mortality rates. Taken collectively with the aforementioned publications, our results provide reassurance that the number of death reports in VAERS for COVID-19 vaccines does not signify an increased risk of mortality with vaccination. Our results add to the evidence base supporting safe use of COVID-19 vaccines and may help reduce vaccine hesitancy.

## Data Availability

VAERS data are available to the public online at

https://vaers.hhs.gov/data.html

## Acknowledgments

We wish to acknowledge the following individuals: Arthur Presnetsov (Centers for Disease Control and Prevention) for querying and formatting of the vaccine administration data; Melvyn Okeke (Food and Drug Administration) for direct informatics support during the analysis.

