## Supplemental Table 1 for "Reporting Rates for VAERS Death Reports Following COVID-19 Vaccination, December 14, 2020-November 17, 2021"

Table S1. Selected VAERS Consolidation Rules for Death Events Query

| VAERS Element | Consolidation Rule |
|---|---|
| Consolidated Age | Derived from the VAERS report associated with the following Reporter Type (by order of priority): Provider, Patient/parent/guardian/caregiver, Manufacturer, Other, Unknown |
| Consolidated Gender | In VAERS reports with conflicting information on gender (male and female in different reports), the derived value is “M/F,” and 'Unknown' values are disregarded.* |
| Consolidated Flag Died | If at least one of the associated VAERS reports has an outcome of “Patient died” selected in Box 21 of the VAERS reporting form, then the Consolidated value is set to ‘Yes’. Otherwise, the value is set to ‘No’. |
| Consolidated Vaccine Date | Derived from the VAERS report associated with the following Reporter Type (by order of priority): Provider, Patient/parent/guardian/caregiver, Manufacturer, Other, Unknown |
| Consolidated Onset Date | Derived from the VAERS report associated with the following Reporter Type (by order of priority): Provider, Patient/parent/guardian/caregiver, Manufacturer, Other, Unknown |
| Consolidated Onset Interval | Derived by subtracting (in days) the Consolidated Vaccine Date from the Consolidated Onset Date |

*Manual review was performed in six cases with conflicting information on gender. In these cases, the narrative text and/or medical records were used to adjudicate gender.
